# Black EquaLity in OCD NeuroGenomics (BELONG): Study Protocol

**DOI:** 10.64898/2026.01.08.26343678

**Authors:** Iasha J. Williams, Dalia Y. Marquez, Kara E. Lopez-lengowski, Malini Bommiasamy, Ogechi Cynthia Onyeka, Slayton J. Underwood, Juliana E. Avery, Jacob Gluckman, Thariana Pichardo, Rhea Chandler, Yulu Brown, Katie Mangen, Emma Grace Choplin, Black EquaLity in OCD NeuroGenomics (BELONG) Study Team, Sonyia C. Richardson, Joseph D. Buxbaum, Eric A. Storch, James J. Crowley, Sidney H. Hankerson, Dorothy E. Grice

## Abstract

Obsessive-compulsive disorder (OCD) is a chronic, serious psychiatric disorder that affects 2-3% of the population and is associated with high personal and societal costs. Genetic factors are estimated to explain roughly half the risk of developing OCD, and genomic studies are just beginning to identify common and rare genetic variants mediating this risk. A major goal of genomic studies is to yield insights into the etiology of OCD and identify molecular targets for the development of novel therapeutics. However, the overwhelming majority of subjects in existing genetic studies are of European ancestry, limiting the generalizability of these findings. To address this gap in understanding, we established the Black EquaLity in OCD NeuroGenomics (BELONG) study (https://belongocd.com/). BELONG aims to collect DNA and clinical data from 1,250 richly phenotyped OCD cases of African ancestry in a culturally sensitive manner. In addition, BELONG includes the collection of parental DNA samples for trio-based analyses and unrelated matched controls for case-control analysis. DNA samples will be sequenced using optimized approaches that will allow us to examine both rare and common genome-wide variation to identify OCD risk genes. We will also meta-analyze these data with other existing OCD genomic data. Overall, BELONG will increase the representation of Black Americans in OCD genetic research, which is necessary to generalize precision medicine discoveries in psychiatric genetics.

## 1 RATIONALE

### 1.1 Overview

OCD is often a lifelong and serious psychiatric disorder affecting 2-3% of the population and is associated with high personal and societal costs.[Fontenelle et al., 2006; Ruscio et al., 2010; Hirschtritt et al., 2017; Stein et al., 2019; Markarian et al., 2010; Lack et al., 2009] OCD can onset throughout the lifespan and significantly impact both childhood and adulthood, impacting developmental tasks, daily functioning, family life, educational and vocational achievements, and overall health.[Lenhard et al., 2021] Impairments associated with OCD can be substantial, and although evidence-based treatments exist and are effective in improving symptom burden and quality of life, a substantial proportion of patients do not fully respond to first-line treatments.[Bloch and Storch, 2015; McGuire et al., 2012] High rates of comorbidity are also associated with OCD;[Torres et al., 2016; Mahjani et al., 2022] for example, the National Comorbidity Survey Replication shows that 90% of individuals with a lifetime OCD diagnosis have at least one additional lifetime psychiatric condition.[Ruscio et al., 2010] OCD is also associated with an elevated risk of co-occurring chronic tic disorders (CTD)[Browne et al., 2015; Mahjani et al., 2022] and other OCD-related disorders. Furthermore, a central role for genetic factors in risk for OCD has been well-established through twin, family, and large cohort studies.[Taylor, 2011; Browne et al., 2015; Mahjani et al., 2021a]

Most large-scale genomic discoveries are heavily biased towards individuals of European ancestry (EUR), despite the fact that EUR populations only make up 16% of the global population, potentially exacerbating existing health disparities.[Martin et al., 2019; Peterson et al., 2019] Samples from diverse ancestries present both challenges and opportunities in genomics research.[Hindorff et al., 2018; Wojcik et al., 2019; Peterson et al., 2019] Integrating samples of diverse ancestry include addressing potential differences in allele frequencies, effect sizes, and linkage disequilibrium (LD) patterns across populations. However, the opportunities include: 1) increasing equity across populations and access to ancestry-informed clinical information; 2) facilitating the identification of additional risk loci[Jun et al., 2017] while also gathering information about high risk gene variants in under-represented populations;[Reitz et al., 2013; Petrovski and Goldstein, 2016; Kunkle et al., 2021] 3) enhancing the fine mapping of genome-wide association study (GWAS) loci through cross-ancestry analysis;[Wojcik et al., 2019] 4) constructing polygenic risk scores (PRS) from cross-ancestry meta-analysis with improved overall prediction, especially for the lesser-represented ancestry[Márquez-Luna et al., 2017; Grinde et al., 2019] and, 5) optimizing actionable variants, which, even for well-established pharmacogenetic risk genes, are still not optimized for diverse populations and likely contribute to perceptions of reduced value of some genetic testing.[Suarez-Kurtz, 2021]

The Psychiatric Genomics Consortium (PGC) OCD Working Group has significantly advanced OCD genomics by recently publishing the first well-powered OCD GWAS, with a sample comprised of over 50,000 cases and approximately 2 million controls.[Strom et al., 2024] However, almost all participants were of EUR ancestry, and phenotypic quality was variable. Recruiting OCD samples from diverse ancestries is essential for advancing OCD gene discovery and ensuring applicability of genetic discoveries to all people with OCD. At least one other study is addressing this gap, actively recruiting 5,000 Latin American individuals with OCD for the NIH-funded Latin American Trans-ancestry INitiative for OCD genomics (LATINO).[Crowley et al., 2023]

In contrast, Black American individuals with OCD are not systematically studied in genetic analyses, a key gap that this study aims to address. In fact, Black Americans with OCD are vastly underrepresented, or altogether absent, from most research studies of any kind, despite the evidence for 1) disparities in access to clinical care, including low rates of participation in specialty treatment, 2) persistent OCD due to lack of treatment, 3) differences in OCD subtypes in Black American populations, as well as 4) the amplification of mental health disparities during the COVID-19 pandemic (including OCD and anxiety disorders) in this population.[Williams et al., 2012a; Himle et al., 2008; Brooks et al., 2022; Williams et al., 2017] These disparities highlight long-standing inequities in education, income, and other socio-demographic factors linked to structural racism.[Moise and Hankerson, 2021; Hankerson et al., 2022] In addition to more structural-level analyses, it is critical to address cultural and contextual factors influencing OCD diagnoses and research participation among Black Americans. These factors include mistrust of researchers, mental health stigma, reliance on faith or individually-based outlets for emotional support, as well as lower rates of health literacy, likely resulting from the aforementioned factors.[Hankerson et al., 2015; Svob et al., 2023] Failing to account for these factors may perpetuate the current status quo in which Black Americans with mental health conditions such as OCD who seek treatment are more likely to be misdiagnosed,[Chasson et al., 2017] terminate treatment prematurely, or be hospitalized.[Hankerson et al., 2011]

We have designed the Black EquaLity in OCD NeuroGenomics (BELONG) study (https://belongocd.com/) to address the urgent need to include Black Americans in OCD genomics (see ***Figure 1***). This is a multi-site study composed of clinical researchers, geneticists, and neuroscientists, with expertise in assessing OCD, collecting biospecimens, conducting genetic analyses and implementing community partnership approaches. One focus of this study is rare genetic variation, variation that carries a high effect size and are often de novo. As such, recruitment prioritizes participants who have OCD and whose biological parents are also willing to participate, since parental genomes serve as ideal controls when studying de novo genetic variation. Unrelated controls will also be recruited to support the case-control study design. Future genetic and clinical analyses will also provide insight into common genetic variation in OCD risk and additional understanding regarding the phenotypic and clinical trajectories of OCD and social determinants of health in this cohort of Black Americans.

**Figure 1.**
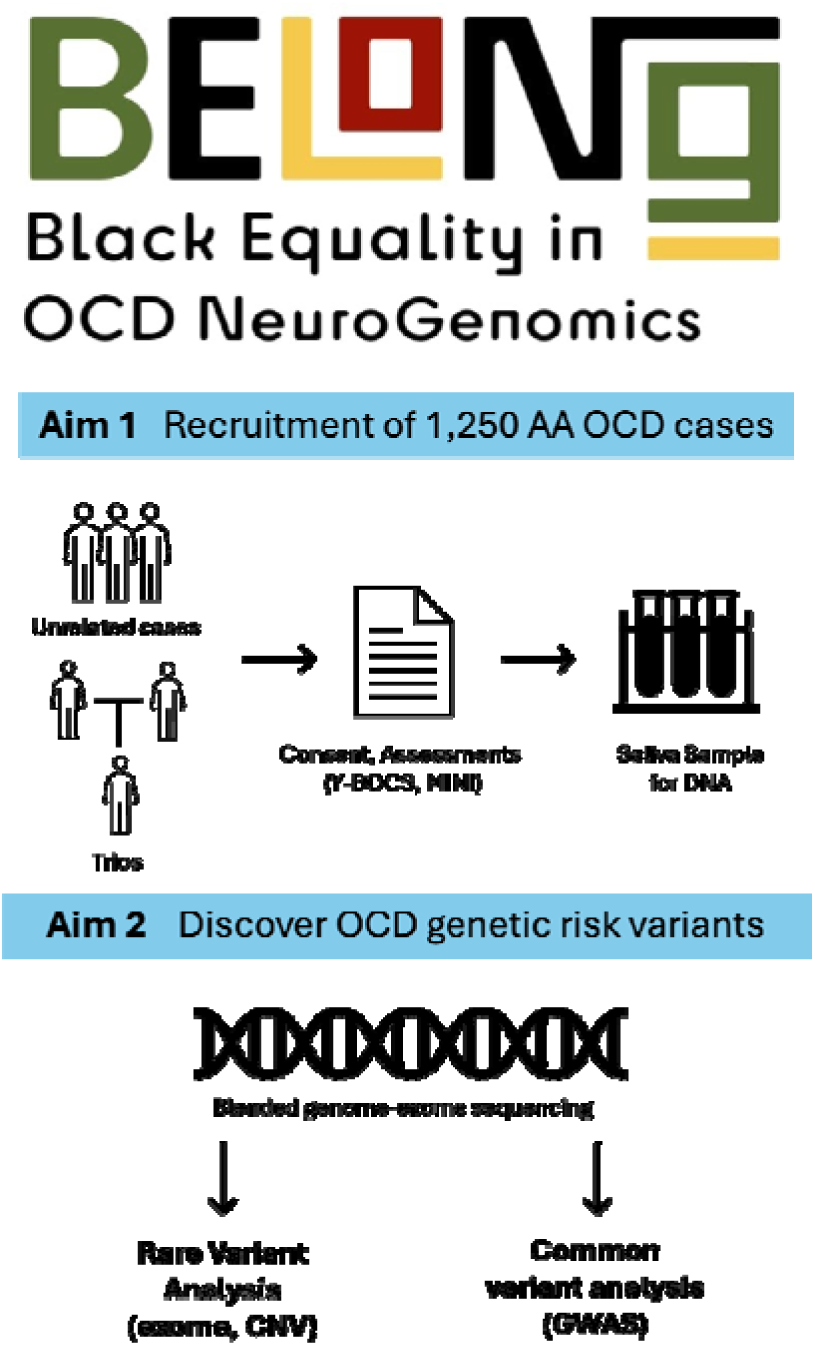
Overview of study aims and protocol.

### 1.2 Background

OCD is a neuropsychiatric disorder characterized by recurrent and intrusive unwanted thoughts, ideas, images, or urges (known as obsessions) and repetitive ritualized behaviors or mental acts (known as compulsions) that are typically performed in response to obsessions.[American Psychiatric Association, 2013; World Health Organization, 1992] Individuals with OCD experience exaggerated concerns and interfering anxiety about a broad range of themes, often related to danger, hygiene, responsibility, or harm, resulting in persistent conscious attention to the perceived threat (obsessions) and distressing preoccupations.[Goodman et al., 2021] In response to this distress, the individual performs compulsions which serve to neutralize their distress and provide some temporary relief. However, in OCD, the obsessions inevitably return and since the relief provided by compulsions is negatively reinforcing, this leads to ongoing, repetitive, compulsive behaviors when obsessions recur.[Pauls et al., 2014] This multidimensional disorder consists of roughly four primary symptom dimensions, including contamination, symmetry, forbidden thoughts and related compulsions, and checking,[Bloch et al., 2008] which may have distinct neural circuitry,[Mataix-Cols et al., 2004] genetic,[Hasler et al., 2007] and etiological origins.[Leckman et al., 2009] Additionally, some subtypes exist, such as OCD that onsets prior to puberty (“early onset”) and tic-related OCD, which occurs when an individual meets diagnostic criteria for OCD as well as for a tic disorder, may be etiologically meaningful.[Brander et al., 2019; Leckman et al., 2010]

Gaining a deeper understanding of the pathophysiology of OCD is essential for developing more effective treatments. Although medications and specialized behavioral therapies are often beneficial and reduce symptom severity and degree of interference,[McGuire et al., 2015; Olatunji et al., 2013] symptom control remains imperfect, and the disorder typically follows a chronic intermittent course. This study of OCD with its focus on Black Americans, who face similar prevalences of OCD to the overall global population, [Himle et al., 2008; Ruscio et al., 2010; Zhang and Snowden, 1999] but are poorly represented in most OCD research studies, will contribute to our understanding of the etiology of this disorder, and include an understudied population. Identifying gene variants (and then the related biological pathways and neurocircuitry) that confer high risk for OCD will help for the basis of new, more specific treatments and an improved understanding of specific phenotypic or clinical differences in this cohort will help to enhance treatment strategies, differential diagnosis and detection methods.[Grassi et al., 2020]

### 1.3 OCD Genetics

Twin and family studies first demonstrated a significant genetic component to OCD, with heritability estimates of ∼30-40%.[Taylor, 2011; Browne et al., 2015; Mahjani et al., 2021a] Work by Grice and team in a national cohort of 822,843 individuals provided heritability estimates of 32-35%, as well as evidence for a significant role of maternal effect (also termed “maternal genetic nurture”) in OCD risk.[Mahjani et al., 2020] Both common and rare genetic variation contribute to the risk for OCD. Regarding common variation, the Psychiatric Genomics Consortium (PGC) OCD Working Group recently published an OCD GWAS with 53,660 cases and ∼2 million controls.[Strom et al., 2025] This GWAS is a significant step forward in OCD genomics, revealing the first 30 genome-wide significant loci for OCD. The identified risk loci are beginning to shed light on biological mechanisms at play in OCD. For example, a novel association in the major histocompatibility region of chromosome 6 is of particular interest because autoimmunity has been repeatedly linked to OCD in genetic and epidemiological studies.[Mataix-Cols et al., 2018; Tylee et al., 2018; Westwell-Roper et al., 2019] This new GWAS is an important advance and is facilitating informative “post-GWAS” studies, such as polygenic risk score analyses.

Regarding rare variation, studies of copy number variation (CNV) were the first to support a role for rare deleterious variation in OCD genetic risk.[McGrath et al., 2014; Gazzellone et al., 2016; Zarrei et al., 2019; Mahjani et al., 2021b] Sequencing studies have also provided strong support for rare single nucleotide variation (SNV) in the risk for OCD, including de novo mutations.[Cappi et al., 2020; Halvorsen et al., 2021] Cappi et al. observed that rates of de novo mutations, likely protein-truncating variations (PTV) (i.e., creation or loss of a stop codon, disruption of a canonical splice site, or a frameshift indel), are significantly elevated in OCD trios.[Cappi et al., 2020] Similarly, Halvorsen et al. observed a 1.3-fold increase in damaging de novo variants in OCD cases relative to controls, including a 2.6-fold increase in de novo PTV in constrained genes.[Halvorsen et al., 2021] While these results were based on analysis of small samples, Cappi et al. identified CHD8 and SCUBE1 as high-confidence OCD genes, with significant overlap of findings with CTD and autism studies and Halvorsen et al. identified CHD8, CUL3, COL4A3, FN1, and ADIPOR2 as genes of interest from their trio data analysis. Using the Transmitted And *De novo* Association (TADA) association test to analyze these two datasets, along with additional OCD samples sequenced by our group, indicates that CHD8 is a genome-wide significant finding in OCD (manuscript in preparation).

## 2 AIMS AND METHODS

### 2.1 Aim 1

#### 2.1.1 Recruitment

The Icahn School of Medicine at Mount Sinai, Baylor College of Medicine, and the University of North Carolina-Chapel Hill maintain strong ties to their surrounding communities in New York City, Houston, and Chapel Hill, respectively, and have previously demonstrated excellence in research and/or clinical treatment services, allowing the ability to recruit a large cohort of research participants. Methods of recruitment include disseminating physical flyers, social media, advertising at relevant conferences and community events, and the use of electronic health records (EHR). We have found the combination of physical and digital advertising and EHR-based ascertainment to be robust and cost-effective recruitment strategies for these types of studies.

#### 2.1.2 Participants

To meet inclusion criteria, OCD case participants must fulfill the diagnostic criteria for OCD via ICD-10 and are included in the study regardless of other psychiatric comorbidities. Individuals are excluded in cases of diagnostic uncertainty, such as OCD secondary to a neurological disorder, or when the differential diagnosis between OCD and another condition is unclear.

Participants must identify as Black or African American and/or have at least one parent or grandparent who identifies as Black or African American.

Parents of confirmed OCD cases (to compose a trio) represent important genetic controls. Unrelated individuals who do not meet the diagnostic criteria for OCD via ICD-10 are population controls and are included in the study regardless of non-OCD psychiatric comorbidity. Unrelated controls must also self-report as Black or African American, and/or have at least one parent or grandparent who identifies as Black or African American.

#### 2.1.3 Approvals and informed consent

All sites have received approval from their institutional IRB to perform this study. All participants provide their informed consent to enter this study, either in writing or via approved electronic consent, including a video call consent process with a study team member. Participants receive modest compensation upon completion of the initial assessment and a second time after providing a saliva sample. Consents for both cases and controls include permission to be recontacted and to share de-identified genetic and phenotypic data with authorized researchers.

#### 2.1.4 Biomaterial collection for DNA analysis

All participants donate one tablespoon (∼15 ml) of saliva for DNA analysis using standardized collection kits. Participants are instructed to refrain from eating, drinking, smoking, or chewing gum for 30 minutes prior to collecting the sample. Participants recruited in person are provided with a saliva kit for sampling during their appointment, while those recruited online receive a saliva kit by mail with a prepaid return label. DNA from 15 ml of saliva is extracted using standard automated processes.

#### 2.1.5 Clinical assessments

The full assessment battery (see ***Table 1***) includes a combination of measures that are clinician-administered, self-report, or parent-report (for participants under the age of 18).

**Table 1.**
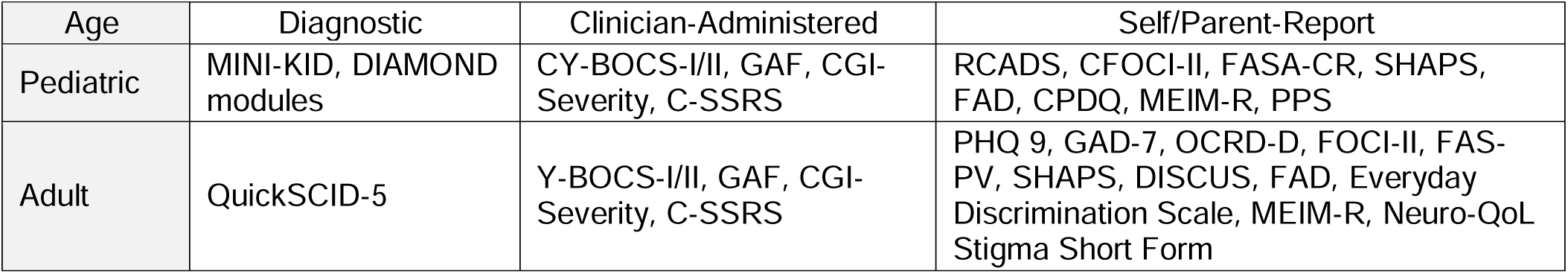
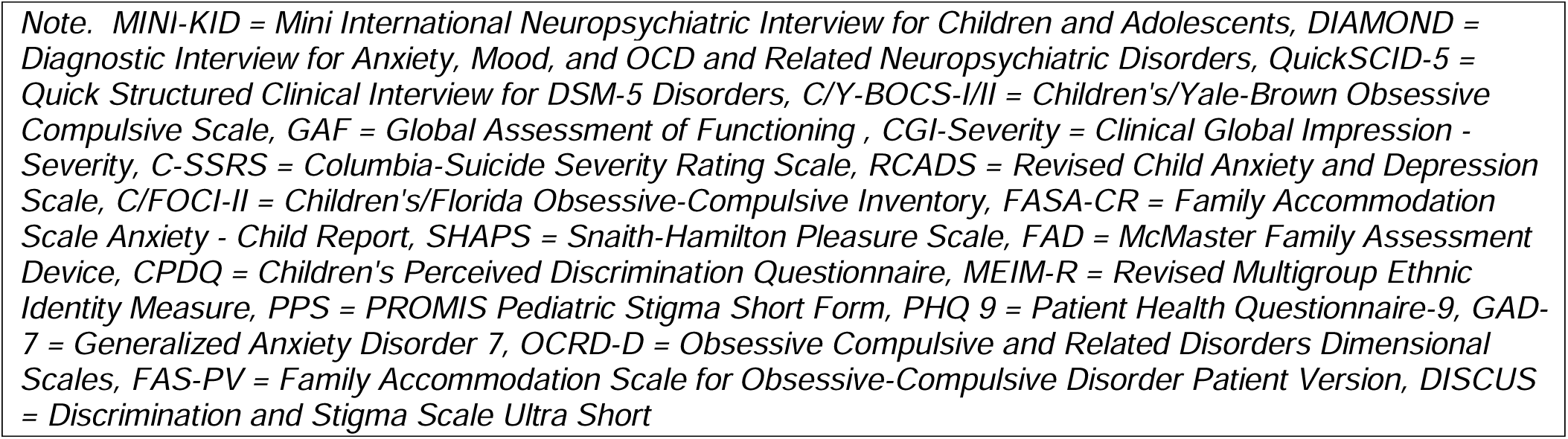
Harmonized OCD diagnostic, treatment and outcome measures to be collected.

| Age | Diagnostic | Clinician-Administered | Self/Parent-Report |
| --- | --- | --- | --- |
| Pediatric | MINI-KID, DIAMOND modules | CY-BOCS-I/II, GAF, CGI-Severity, C-SSRS | RCADS, CFOCI-II, FASA-CR, SHAPS, FAD, CPDQ, MEIM-R, PPS |
| Adult | QuickSCID-5 | Y-BOCS-I/II, GAF, CGI-Severity, C-SSRS | PHQ 9, GAD-7, OCRD-D, FOCI-II, FAS-PV, SHAPS, DISCUS, FAD, Everyday Discrimination Scale, MEIM-R, Neuro-QoL Stigma Short Form |

*Note. MINI-KID = Mini International Neuropsychiatric Interview for Children and Adolescents, DIAMOND = Diagnostic Interview for Anxiety, Mood, and OCD and Related Neuropsychiatric Disorders, QuickSCID-5 = Quick Structured Clinical Interview for DSM-5 Disorders, C/Y-BOCS-I/II = Children's/Yale-Brown Obsessive Compulsive Scale, GAF = Global Assessment of Functioning, CGI-Severity = Clinical Global Impression - Severity, C-SSRS = Columbia-Suicide Severity Rating Scale, RCADS = Revised Child Anxiety and Depression Scale, C/FOCI-II = Children's/Florida Obsessive-Compulsive Inventory, FASA-CR = Family Accommodation Scale Anxiety - Child Report, SHAPS = Snaith-Hamilton Pleasure Scale, FAD = McMaster Family Assessment Device, CPDQ = Children's Perceived Discrimination Questionnaire, MEIM-R = Revised Multigroup Ethnic Identity Measure, PPS = PROMIS Pediatric Stigma Short Form, PHQ 9 = Patient Health Questionnaire-9, GAD-7 = Generalized Anxiety Disorder 7, OCRD-D = Obsessive Compulsive and Related Disorders Dimensional Scales, FAS-PV = Family Accommodation Scale for Obsessive-Compulsive Disorder Patient Version, DISCUS = Discrimination and Stigma Scale Ultra Short*

Participants, whether cases or controls, complete the same assessment battery tailored for either adults or youth. Parent participants provide demographic information instead of a full assessment.

#### 2.1.6 Clinician-administered measures

The Quick Structured Clinical Interview for DSM-5 Disorders (QuickSCID-5)[First and Williams, 2020] and the Mini International Neuropsychiatric Interview for Children and Adolescents (MINI-KID)[Sheehan et al., 2010] are structured diagnostic interviews to assess DSM-5 and ICD-10 disorders in adults and youth, respectively. Select modules of the Diagnostic Interview for Anxiety, Mood, and OCD and Related Neuropsychiatric Disorders (DIAMOND)[Tolin et al., 2018] are administered for children and adolescents to evaluate obsessive-compulsive related disorders or OCRDs. Both the Yale-Brown Obsessive Compulsive Scale (Y-BOCS-I/II)[Goodman et al., 1989; Storch et al., 2010] and the Children’s Yale-Brown Obsessive Compulsive Scale (CY-BOCS-I/II)[Scahill et al., 1997; Storch et al., 2019] are semi-structured interviews used to assess the presence of distinct obsessive-compulsive symptoms and overall severity. To evaluate suicidal ideation, risk factors, and behaviors, the Columbia-Suicide Severity Rating Scale (C-SSRS)[Posner et al., 2007, 2011] is administered by raters. Following the administration of these comprehensive measures, raters complete the Global Assessment of Functioning (GAF)[Schwartz and Del Prete-Brown, 2003] and the Clinical Global Impression - Severity (CGI-S)[Guy, 1976] to assess overall psychological and social functioning and impairment.

#### 2.1.7 Self/Parent-report measures

Assessment visits with adult participants include completion of self-report surveys regarding information on psychiatric symptoms and participants’ quality of life. The Patient Health Questionnaire-9 (PHQ-9)[Kroenke et al., 2001] and Generalized Anxiety Disorder 7 (GAD-7 Anxiety)[Spitzer et al., 2006] assess symptoms of depression and anxiety, respectively. The Obsessive-Compulsive and Related Disorder Dimensional Scales (OCRD-D) [LeBeau et al., 2013] gathers information on OCRDs in adults. The Florida Obsessive-Compulsive Inventory (FOCI-II)[Storch et al., 2007] collects further self-reported data on OCD symptoms and severity. Participants’ families’ accommodation of their OCD symptoms is measured by the Family Accommodation Scale for Obsessive-Compulsive Disorder Patient Version (FAS-PV).[Wu et al., 2016] An abbreviated adaptation of the Discrimination and Stigma Scale (DISC-12),[Thornicroft et al., 2009] the Discrimination and Stigma Scale Ultra Short (DISCUS),[Bakolis et al., 2019] and the Everyday Discrimination Scale[Williams et al., 1997] are employed to assess perceived discrimination and stigma related to OCD and other personal characteristics, respectively. Several self-report measures overlap between adult and youth assessments to further evaluate elements of quality of life. The Snaith-Hamilton Pleasure Scale (SHAPS)[Snaith et al., 1995] consists of questions aimed at understanding participants’ hedonic experiences. Items in the Revised Multigroup Ethnic Identity Measure (MEIM-R)[Phinney and Ong, 2007] gather information on participants’ views of their ethnic group, while the McMaster Family Assessment Device (FAD)[Epstein et al., 1983] expands on participants’ perceptions of their familial dynamics. The Neuro-Qol Stigma Short Form for Adults is an 8-item bank measuring the perceived impact of stigma on quality of life for individuals with OCD (Quality of Life in Neurological Disorders).[Cella et al., 2012]

Youth assessments feature three additional measures that differ from adult assessments. The Revised Child Anxiety and Depression Scale (RCADS)[Chorpita et al., 2000, 2005] is a self-report questionnaire evaluating anxiety and depression symptoms in youth. The Family Accommodation Scale Anxiety – Child Report (FASA-CR)[Lebowitz et al., 2013, 2015] uses modified language for youth to assess family accommodation of participants’ anxiety symptoms. Items from the Children’s Perceived Discrimination Questionnaire (CPDQ)[LaFont et al., 2018] gather information on youth participants’ perceptions of discrimination experiences. The PROMIS Pediatric Stigma Short Form (PPS)[Lai et al., 2024] is a youth-focused measure evaluating perceived stigma experiences related to OCD symptoms.

#### 2.1.8 Demographic assessments

In-depth demographic information is collected, ranging from core data (e.g., age, sex, education, etc.) to additional diagnoses and treatment history. Information on the perceived efficacy of previously received treatments (e.g., psychotherapy, medication, etc.), access to treatment, and factors contributing to symptom improvement are all assessed. Data are collected on medications used for treatment, including whether they are current, the highest dose, duration, primary reason for prescription, and impact on OCD symptoms. Written responses regarding participants’ views on how OCD treatments might be improved, as well as their beliefs about how OCD has impacted their quality of life, are also gathered.

#### 2.1.9 Clinical data collection, quality assurance, and analysis

The HIPAA-compliant Research Electronic Data Capture (REDCap) digital database, direct data entry platform, and survey collection tool are utilized to collect phenotypic data.[Harris et al., 2009, 2019] Data quality is assessed through REDCap-enabled automatic alerts, manual data checks consistently performed by each site, and monthly cross-site consultations to ensure data harmonization and inter-rater reliability (IRR). IRR meetings are facilitated by a licensed clinical psychologist with expertise in evaluating OCD in Black patients. Issues of missing or incorrectly entered data are communicated to the coordinating site, and any necessary steps are taken to correct current issues and mitigate any future errors. Study visits are audio/video recorded according to participant preference to further assess IRR using *Zoom* (version 6.3.10) and *Microsoft Teams*. Local site PIs provide weekly (C)YBOCS I-II, QuickSCID-5, and MINI-KID supervision to evaluate the accuracy and quality of assessments.

Clinical and demographic data captured will be analyzed to update and expand the current literature on OCD in Black Americans. Analysis of data gathered from the clinician-administered measures will contribute to existing knowledge on differences in obsessive-compulsive symptom presentation and common comorbidities among Black Americans both with and without OCD. Clinical interview data are expected to replicate previous findings that Black Americans with OCD report greater contamination, perfectionism, and fears of being misunderstood concerns[Williams et al., 2012b]. Self-report surveys, in turn, will provide deeper insight into previously demonstrated obstacles for Black Americans with OCD such as delays in diagnosis, limited access to treatment, and discrimination.[MacIntyre et al., 2023; Williams et al., 2012a] Collectively, all data gathered will be analyzed and disseminated to fill the current gaps in this area of research. These gaps include longstanding underrepresentation of Black participants in OCD research, a dearth in research examining both the clinical and genetic aspects of OCD in this population, and a lack of studies looking at OCD in Black youth.[Williams et al., 2012b; Williams and Jahn, 2017]

### 2.2 Aim 2

#### 2.2.1 Genetic analysis

We will be capturing both common and rare genetic variation in this study as described in detail below. We employ current best practices for these analyses, which may be updated as methods and technologies advance.

Blended Genome-Exome (BGE) sequencing has recently been developed as a cost-effective approach to common variation across the genome and rare coding variation, replacing both genotype microarrays and whole exome sequencing (WES).[DeFelice et al., 2024] In brief, a whole genome library is generated, from which an aliquot is exome-enriched. The two libraries are then combined (33% exome, 67% genome) into a single tube for sequencing, yielding a single file with a low-coverage whole genome (2-3x) plus a high-coverage exome (30-40x). BGE data allows the imputation of common variants across the genome without the bias introduced by genotyping arrays, which are limited to known variants. This is particularly important in BELONG, as Black American reference panels are smaller than their EUR counterparts. Furthermore, BGE yields high-coverage exome sequencing data, improving confidence in calling rare coding variations. Given that BGE is currently cost-effective compared to WGS, we are using this approach.

Variant calling follows the Genome Analysis Toolkit (GATK) best practices to jointly call single nucleotide variants (SNVs), insertion-deletions (indels), and copy number variants (CNVs). Sequencing reads are aligned to the human reference genome GRCh38 using bwa-mem, marking duplicate reads and conducting base quality score recalibration using Picard. CNVs are called from the deep coverage exome capture loci using GATK-gCNV, which adjusts for known Whole Exome Sequencing (WES) read-depth confounders (e.g., GC content, mappability), while also adjusting for technical batch confounders.

For quality control, we employ a multi-step procedure to ensure that BGE data are of high quality. This includes 1) filtering samples with low WES coverage (<90% of the exome at 10x) or low WGS coverage (mean < 1x), 2) removing samples with chimeric or contamination read rates >5%, 3) excluding samples with discrepancies between imputed genetic sex and reported sex, 4) eliminating closely related individuals by dropping one member of each pair with a kinship coefficient (pi-hat) >0.2 (second degree or closer relatives), while keeping the sample with the best quality control metrics, and 5) for exome analyses, filtering exonic variants with any of the following features: outside of Twist target capture regions, located in a low-complexity genomic region, read depth < 10 x, genotype quality <20, allele balance < 0.0.2 or > 0.0.8 in heterozygous calls, or allele balance < 0.0.8 in homozygous alternate calls.

#### 2.2.2 Rare Variant analyses

First, variants are annotated with the Variant Effect Predictor (VEP), prioritizing coding canonical transcripts. Variants are classified as PTV if annotated by VEP as nonsense, frameshift, or affecting an essential splice site. Prior work in EUR individuals has shown that de novo PTV occur in OCD subjects more frequently than expected by chance and at a higher rate than de novo VEP-annotated missense variants. PTV also exhibit differential enrichment based on measures of functional severity that assess evolutionary constraint against deleterious genetic variation (e.g., pLI or LOEUF scores). For variants annotated as missense by VEP, we will use contemporary methods to delineate classes of variant missense based on putative functional effect.

Second, for gene discovery, we will use the best approaches available at the time of analysis. At present, the transmitted and de novo (TADA) model[He et al., 2013] allows one to simultaneously analyze de novo, transmitted, and case-control rare variant data. TADA is a Bayesian model that first computes a gene-specific Bayes factor for each mutation category and inheritance pattern, and then multiplies these Bayes factors to generate a statistic that summarizes all evidence of association for each gene. The total Bayes factor is ultimately converted into a q-value to control the false discovery rate (FDR).[Rubeis et al., 2014; Sanders et al., 2015; Satterstrom et al., 2020] We will incorporate de novo variation, inherited variation, and case-control variation, including robust Black American control data (if needed, variant calls in Black American samples in gnomAD). Analyses will be performed first on the BELONG cohort, and results from these analyses will be compared to published and unpublished data from other populations. Finally, assuming the mutational patterns are similar across populations, results from all studies will be combined to enhance OCD gene discovery.

#### 2.2.3 GWAS

First, we will impute common variants using the most appropriate reference panel available at the time of analysis. Second, we will perform association testing within BELONG using methods that account for genetic admixture, which we expect in a Black American population. Admixed individuals possess DNA with more than one ancestral component and have been systematically excluded from genetic studies due to the historical lack of available tools to appropriately handle their complex genomes. If not accounted for, such population substructure can infiltrate analyses and bias results when admixed samples are included alongside more homogeneous individuals in standard analysis.[Sul et al., 2018] To conduct gene discovery in the best-calibrated manner for Black American populations, we will utilize methods such as *Tractor*, which corrects for fine-scale population structure at the haplotype level by using local ancestry, allowing admixed samples of all mixture proportions to be readily included in large-scale collections. *Tractor* has been shown to boost GWAS power in admixed cohorts to detect ancestry-enriched loci that may have been undiscoverable in prior European GWAS.

Third, we will perform the largest and most diverse OCD GWAS possible by meta-analyzing BELONG data with all available worldwide OCD GWAS data. This trans-ancestry meta-analysis will include data from PGC OCD, the LATINO study, and any other studies with relevant data. To avoid technical biases introduced by comparing sequencing and array data, we will meta-analyze a common set of variants that are well-imputed in PGC OCD, LATINO, and other relevant studies. We will run GWAS using methods like the Scalable and Accurate Implementation of Generalized mixed model package, SAIGE,[Zhou et al., 2018], which runs a linear or logistic mixed model including a kinship matrix as a random effect and covariates as fixed effects. Methods of this sort account for heterogeneity in sample makeup and provide accurate P values when case-control ratios are imbalanced. To declare genome-wide significance, we will adhere to a P value threshold of 5×10-8.[Pe’er et al., 2008]

## 3 DISCUSSION

While the field of psychiatric genetics is advancing at a notable speed, OCD genetic studies have lagged behind those of other disorders.[Grice, 2020] Therefore, the recent PGC OCD GWAS, which describes the first 30 genome-wide significant loci for OCD, represents a major advance in our field. However, as noted previously, the PGC OCD GWAS lacks genomic diversity among its OCD cases and identifies common genetic variants which would carry small effect, not rare genetic variants of large effect. The BELONG study will be a significant advancement in improving diversity in OCD genetic studies and focuses on identifying variants of large effect. In addition, work in other psychiatric disorders has shown that diversifying a sample can markedly accelerate GWAS locus discovery and increase fine-mapping resolution.

Furthermore, although OCD rare variant sequencing studies have made some progress ([Cappi et al., 2020; Halvorsen et al., 2021; Lin et al., 2022]), they suffer from the same lack of diversity as GWAS studies as well as small sample sizes. Overall, both genetic and non-genetic studies of OCD have included few, if any, Black American participants and the BELONG study specifically ascertains and analyzes OCD biological samples and clinical data from Black American populations. All of the genetic data will be meta-analyzed with existing datasets to increase statistical power and maximize diversity.

BELONG will significantly increase the number of sequenced DNA cases of OCD. To date, there are approximately 1,500 published exome or genome sequenced (primarily EUR) OCD cases, and by the completion of this study, we will have an equivalent number of Black American OCD cases. This is important because it will not only increase statistical power but also increase the generalizability of these findings to those of non-European ancestry. Previous rare variant analyses comparing OCD cases to unaffected individuals have found an increased burden of deleterious mutations in constrained genes, thus providing a means to readily identify OCD risk genes.[Cappi et al., 2020; Halvorsen et al., 2021] One future direction for this study is to sample a greater representation of individuals from the African diaspora both within and outside the United States. This would create a more representative sample of OCD on a global scale and improve resolution for identifying high-confidence OCD risk genes. Additionally, this would enable a deeper understanding of environmental and cultural impacts on OCD symptom presentation. Future studies should also consider gathering additional information on factors known to influence OCD symptom presentation, such as religious affiliation, experiences of racial discrimination, and access to quality treatment.

This study is of importance and relevance to Black Americans with OCD and their families as the field of OCD research continues to advance and impact existing understandings and treatment options for the disorder. While many advancements have been made in OCD research in recent years, these findings remain largely non-generalizable to those of racial/ethnic minority backgrounds.[Wilson and Thayer, 2020] To effectively support those with OCD and their families, studies such as this one will be pivotal in ensuring representation exists at all levels and that the impacts of research reach all who are affected by OCD. Continued efforts toward more representative science are imperative in the field of OCD research and beyond.

## Data Availability

Phenotypic data, DNA sequence data, and scripts available for access by other qualified researchers. The datasets generated and analyzed will be available in the NIMH Data Archive (https://nda.nih.gov/). DNA samples will be available from the NIMH Repository and Genomics Resource (https://www.nimhgenetics.org/).

## ACKNOWLEDGMENTS

The BELONG team acknowledges the contributions of Jacquelyne Wynn for her input on various aspects of the project. We’d also like to thank Catherine Borzym for designing the BELONG study website, and Andrew Storch for designing the BELONG study logo. All BELONG sites thank, in particular, the essential contribution of those with lived experience and their families for participating in BELONG.

## ETHICS APPROVALS

BELONG was approved by the MSSM Institutional Review Board (IRB) (Approval Number: 23-01112) under the single IRB (sIRB) framework. MSSM is the IRB of record for all sites.

## CONSORTIA

*Black EquaLity in OCD NeuroGenomics (BELONG)*

All of the named authors on this manuscript, plus:

John Saunders, MD^1,2^, Joura Njoh^2,3^, Tiffanie Williams-Brookes^4^, Ivy Ruths, PhD^5^, Darlene Davis Goodwine, PhD^6^, Bianca Simmons^7^, Catherine Rast^2,8^, Hannah Moore^2^, Vanessa Zavala Cruz^2^, Grace Tisdale^2^, Gianna Colombo^2^, Megan Dailey^2^, Fre’Dasia Daniels^2,3^, Maya Etta^2,3^, Ella Kim^9,10^, Javan Tate, Sean McCartney^2,3^, Asmita Ahuja^2,11^, Dayan Berrones^2^, Javan Tate^2^, Ayisat Adegbindin^2^, Brian Dang^2^

^1^Harris Health Ben Taub Hospital

^2^Department of Psychiatry and Behavioral Sciences, Baylor College of Medicine

3Department of Psychological Health & Learning Sciences, University of Houston

^4^The Harris Center for Mental Health and IDD

^5^Houston Anxiety and Wellness Center

^6^Aidan Behavioral Health & Consulting

^7^Bianca Simmons, LLC

^8^University of Wisconsin-Milwaukee

^9^Department of Psychiatry, Icahn School of Medicine at Mount Sinai

^10^The Spence School

^11^Rice University

## AUTHOR CONTRIBUTIONS

*Data collection*: IJW, DYM, KEL, MB, OO, SJU, JA, JG, TP, RC, YB, KM, EGC.

*Conceptualization*: DEG, SH, EAS, JDB, JJC. *Funding acquisition*: DEG, SH. *Manuscript preparation*: all.

## ON-LINE RESOURCES

BELONG study website: https://belongocd.com/

